# Incentivizing Multiple Objectives in Active Surveillance for Urban Disease Vectors

**DOI:** 10.1101/2021.01.21.21250245

**Authors:** Claudia Arevalo-Nieto, Justin Sheen, Gianfranco Condori-Luna, Carlos Condori-Pino, Julianna Shinnick, Jennifer K. Peterson, Ricardo Castillo-Neyra, Michael Z. Levy

## Abstract

Large-scale vector control campaigns have successfully reduced infectious disease incidence around the world. In addition to preventing new infections, these campaigns produce a wealth of information about the distribution and density of insect vectors, which can be incorporated into risk maps. These maps can effectively communicate risk map data to technicians on the ground, although encouraging them to use the data remains a challenge. We carried out a series of rolling trials in which we evaluated risk map use under different incentive schemes. Participants in the studies were trained field technicians tasked with house-to-house surveillance for insect vectors of Chagas disease in Arequipa, Peru. A novel incentive scheme based on poker best achieved a dual objective: to encourage technicians to preferentially visit higher-risk houses while surveilling evenly across the search zone. The poker incentive structure may be well-suited to improve entomological surveillance activities and other complex multi-objective tasks.

## MAIN

Vector-borne diseases kill at least 700,000 people each year^1^. A common method for interrupting vector-borne disease (VBD) transmission is to target vector populations, especially blood feeding insects, in human-dominated areas, often through large-scale vector control campaigns. These efforts have successfully reduced VBD incidence^2–4^, and in the process have also produced a wealth of information about vector distribution and density^5^. Such data could be invaluable in guiding subsequent surveillance and control activities, but often remains underutilized.

One avenue for utilizing vector control data is to incorporate them into risk maps. In recent years, risk map technology has advanced rapidly^6–10^; computational power now allows for precise inference over large areas^11–13^, while mobile technology can bring up-to-date predictions directly to surveillance personnel in the field^9,14,15^. Nonetheless, achieving optimal use of the complex spatio-temporal information provided by risk maps still hinges on the behavior of the technicians tasked with the job. Simply generating and delivering a map does not ensure its appropriate use^9^, which can require the end user to be cognizant of multiple types of information at once, in addition to changing their habitual behaviors.

In this study, we asked if incentives could help to bridge the gap between risk map delivery and optimal map use. We investigated this question using risk maps for domestic infestations of the Chagas disease vector *Triatoma infestans* in Arequipa, Peru (pop: ∼1.3 million). Chagas disease is a vector-borne infection caused by the parasite *Trypanosoma cruzi*. The disease is chronic if left untreated, leading to serious cardiac, gastrointestinal, and/or peripheral nervous system morbidity in about 30% of those infected^16^.

Over the past two decades, a large-scale insecticide campaign in Arequipa has significantly reduced the number of Chagas disease vector infestations in the city, resulting in infrequent infestations of households (there are no wild vectors in the region) that are sporadically distributed across the urban landscape. At the height of the campaign, over 30 dedicated field personnel worked on vector control. However, that number has dropped with the prevalence of infestation and presently there are no full-time staff dedicated to Chagas disease vector control. Inspections are conducted by technicians who are responsible for a variety of disease prevention activities, in addition to routine laboratory work [Laura Tamayo, pers. comm.]. Our risk maps provide surveillance personnel with the opportunity to make evidence-based decisions when looking for *T. infestans* in the challenging post-vector control campaign scenario, and ultimately, help to prevent vector re-emergence.

## METHODS

### Overview

We conducted a rolling series of four trials in which we evaluated map use by vector control personnel (hereafter referred to as participants) under different incentive schemes. Each trial took place in a district of Arequipa, Peru that had a history of *T. infestans* infestation (described below).

### Ethics statement

All participants provided written informed consent to participate in the study. The study was approved by the institutional review boards of the University of Pennsylvania (protocol number 824603) and the Universidad Peruana Cayetano Heredia (protocol number 66427).

### Study sites

Trials were conducted in four of Arequipa’s twenty-nine districts: the Socabaya district, the Cayma district, the Jose Luis Bustamante y Rivero (JLByR) district, and the Miraflores district. All districts had a population of at least 75,000 people, and a history of substantial *T. infestans* infestation during the insecticide treatment phase of the vector control campaign between 2007 and 2012. Further demographic and historical information for each district are provided in the supplementary materials.

### Experimental design

Each of the four trials had at least two arms in which we tested different incentive schemes. Trials followed a crossover design in which participants were compared to themselves across arms of the same trial^17^. Participants were assigned to a trial arm for five days (Mon-Fri), working for approximately five hours per day. For logistical reasons (vacations), some individuals did not participate in every trial. Six individuals participated in the first three trials, and nine in the final trial.

In each trial arm, a participant was assigned to surveil a new ‘search zone,’ which were contiguous areas within one of the four districts that contained 400-600 households. This number of households is similar to that which a vector control technician might surveil in a given week. The boundaries of the zones were artificial, (as are the politically-defined districts in which they are located), but they generally followed natural features of the landscape. Each search zone was included in the study only once. We randomized the assignment of search zones and the order in which each arm was carried out.

### Risk maps

Risk maps for vector infestation were provided on smart phones using the VectorPoint mobile app (described in detail in^9,18^). Each participant had at least one month of experience using the app prior to the study. Briefly, the VectorPoint app provides a map of each search zone in which each household in the zone is displayed as a dot that is colored one of five colors. Each of the five colors represents the risk of that household being infested with *T. infestans* relative to all other houses in its search zone (Figure 1).

**Figure 1.**
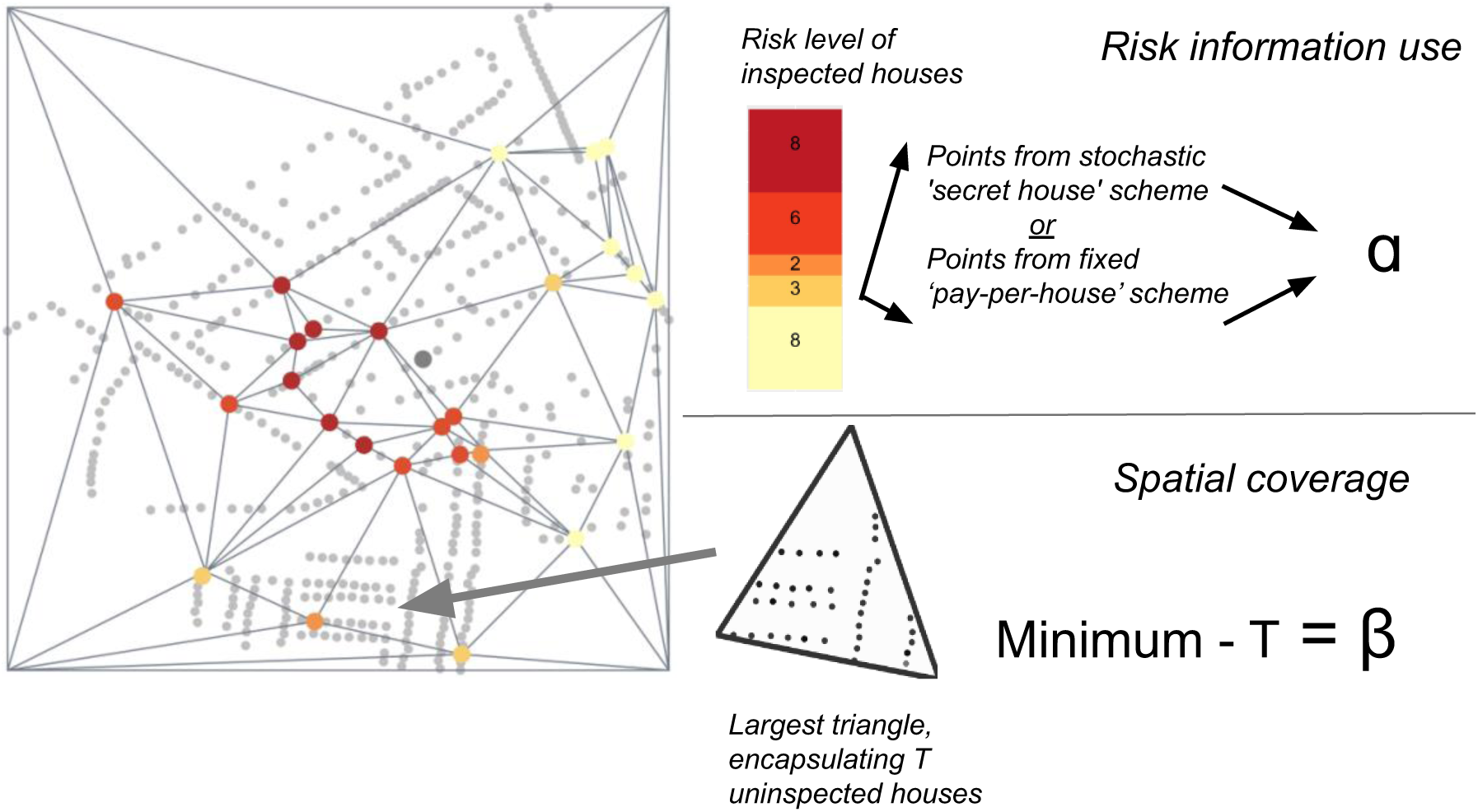
Representation of risk-based and spatial incentives. **Left:** Map displaying a search area of N households, each represented by a dot that is colored one of five colors representing relative risk of vector infestation. Top right: ‘**ɑ’** represents the reward for risk information utilization, which was earned in a stochastic or fixed incentive scheme. Bottom right: **β** represents the reward for spatial coverage, which is calculated by subtracting the maximum number of uninspected houses bounded by Delaunay triangles formed between the inspected houses, T, from the minimum expected coverage (N *.05). In the first three trials, the total reward was a weighted average of **ɑ** and **β**. In the final trial, values of **ɑ** and **β** were added together to form hierarchical ‘poker hands’.

Risk estimates displayed in the map were generated using infestation data from the Arequipa vector control campaign run in a longitudinal, spatio-temporal model described previously^9,18^. The model was run every evening to incorporate new information collected during inspections that day. To avoid any influence from vector surveillance activities in nearby areas, we ran the model on each search zone individually. Risk maps were updated and synched with the app each morning for participant use.

### Spatial coverage

We quantified spatial coverage of the search zone using a new functionality in the VectorPoint app that draws lines between inspected houses and divides the uninspected spaces into triangles. Every time a new house was inspected, the house became a vertex in a set of Delaunay triangles^19^. New triangles were formed in the app in real time, and were immediately visible to the participants, allowing them to monitor their spatial coverage. We used the number of uninspected houses in the largest triangle as the metric with which we evaluated spatial coverage (Figure 1).

### Incentive structures

Each incentive structure was designed to encourage participants to utilize the household-level infestation risk information presented in the risk map while also displaying good spatial coverage of their search zone. We refer to this as our ‘dual objective,’ which we represent visually in Figure 1. Rewards were rewarded in Peruvian soles. One Peruvian sol is equivalent to approximately $0.28USD.

- **Fixed risk information utilization:** Under this incentive scheme, one of five payment amounts was awarded to participants for each house they inspected. The five payment amounts were positively correlated with the five risk levels (as shown by five colors in the risk map) in order to encourage participants to utilize the household level risk information presented in maps.
- **Spatial coverage:** Under the spatial coverage incentive, participants were rewarded for surveilling evenly across the total area of their search zone, quantified as described above. A minimum expected coverage was established as 5% of the total number of houses in each search zone. As a reminder, the inspected houses formed vertices of a set of Delauney triangles^19^. The largest of these triangles--which represents the biggest ‘hole’ in the search area, was identified (Figure 1). If the largest of the triangles contained fewer houses than the minimum expected coverage, a reward was given. The size of the reward increased with each subsequent decrease in the number of households situated in the largest triangle.
- **Secret houses:** With this incentive, we aimed to encourage participants to use the risk information in the maps by providing rewards for visiting certain ‘secret’ houses that were not revealed to participants ahead of time. Participants were informed that houses with a higher infestation risk level had a higher probability of being ‘secret’ houses. We chose secret houses using weighted random selection, in which each house was weighted proportionally to its estimated risk quintile. We informed participants at the end of each day if they had inspected a secret house. Monetary values awarded for inspecting a secret house differed by trial and arm.
- **Pay per detection:** In this incentive structure we aimed to motivate participants to adopt both target strategies by allocating large financial rewards for detecting and inspecting infested houses. The probability of finding an infested house in Arequipa is very low, so the larger reward was balanced by the lower expectation of a payout.
- **Poker incentive structure (Figure 2):** Under this incentive structure points were awarded for achieving the goals described below. When participants accumulated 500 or 1000 points, they could trade the points in for four hours or a day off work, respectively. The possible goals (‘poker hands’) were:

**Figure 2.**
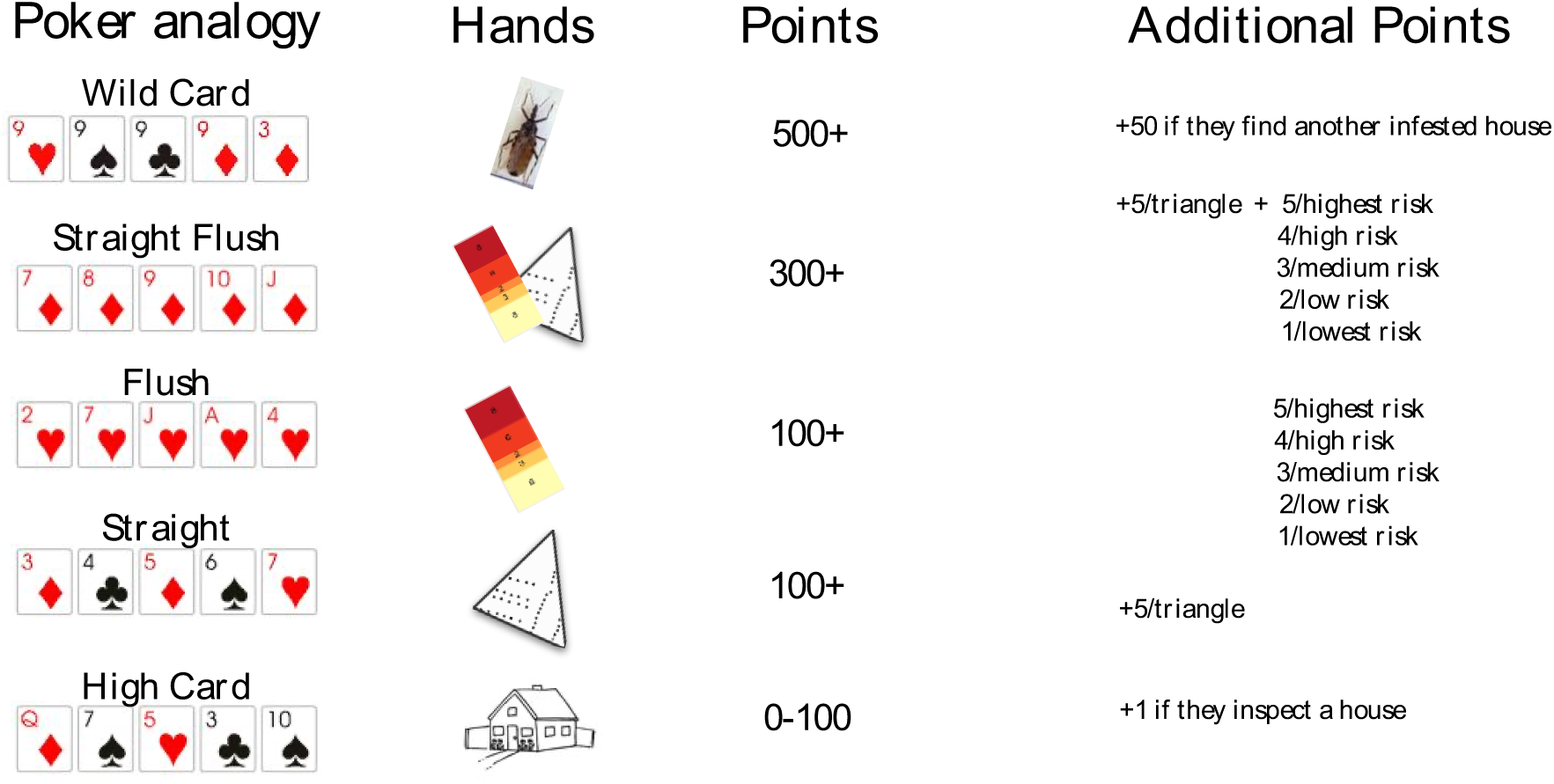
Poker incentive scheme. The first column displays the poker hands corresponding to different ways to earn points. The second column shows the goal that participants could choose to achieve: infested house detection (shown as a bug), higher risk houses (shown as the five-color risk quintile), and good spatial coverage (shown as a map). The house picture represents carrying out home inspections regardless of risk level or spatial coverage. The last two columns describe the base number of points for each hand and additional points that were awarded when participants achieved the goals shown in the second column.
  - **High card (one point per home inspection):** A point was awarded for each home inspection carried out, regardless of spatial coverage achieved or house risk level. A home inspection was analogous to a poker hand that has no flush or straight, and is thus scored on its highest card.
  - **Straight (100 points):** The next level up was a ‘straight,’ which describes a poker hand of five cards in sequence, but of different suits. In this case, spatial coverage was analogous to card sequence; when spatial coverage exceeded 5%, a ‘straight’ was achieved.
  - **Flush (100 points):** A ‘flush’ refers to a poker hand with five cards of the same suit in no sequential order. In our poker incentive scheme, a flush was achieved when the mean risk level of all houses inspected in a week was at least four (out of five possible levels/quintiles).
  - **Straight flush (300 points):** A ‘straight flush’ is a poker hand with five cards of the same suit (the flush) in sequential order (the straight). Hence, a straight flush was achieved under the poker incentive scheme by inspecting houses with a mean risk level of four *and* spatial coverage over 5%.
  - **Wild card (500 points):** Detecting infestations in a low-prevalence situation involves some combination of strategy and luck. We draw the analogy to using a wildcard--which, in some versions of poker, confers great advantages to the player who draws it. In our poker incentive scheme, participants scored a ‘wild card’ by detecting an infested house.

### Statistical analyses

All statistical tests were carried out in the R statistical computing environment^20^. Given the ordinal nature of the outcome variable for risk information utilization, we used a proportional odds logistic regression (POLR function from the MASS Package^21^) to compare the number of houses that participants inspected in each risk quintile under the different arms of each trial. For analyses of spatial coverage in the trials with two arms (Cayma, JLByR and Miraflores trials), we performed a paired t-test for each trial in which we compared spatial between arms. To analyze spatial coverage in the Socabaya trial, which had three experimental arms and one control arm, we carried out a paired t-test comparing spatial coverage in each of the three experimental arms to that of the control arm.

## RESULTS

Table 1 is provided as a quick reference for the incentive scheme descriptions. Trial names reflect the district in which they were carried out. Data for each participant are available in Tables S1 and S2 and in Figure S1. All results are summarized in Table 2.

**Table 1.** Overview of the incentive schemes tested in this study. Detailed information for each scheme is found in the Methods section and Figures 1 and 3.

| Incentive | Description |
| --- | --- |
| Secret houses | Participants rewarded for inspecting houses designated as ‘secret houses’ using random selection weighted toward higher risk houses |
| Spatial coverage | Participants rewarded for decreasing Delaunay triangle size beyond a minimum threshold. |
| Pay-per-detection | Participants rewarded for finding an infested house |
| Poker scheme | Participants rewarded for achieving tasks named for different poker schemes |

**Table 2.** Results overview. ^1^One sol = $0.28 USD. ^2^ Jose Luis Bustamante y Rivero. *Calculated by dividing 0.1,0.2,0.3,0.4, or 0.5 soles by a denominator of 6 soles.

| Trial | Arm | Incentive | Payout | Reward<sup>1</sup> | Behavior change |
| --- | --- | --- | --- | --- | --- |
| Socabaya | A | Secret houses | Stochastic | 4 soles | None |
|  |  | Spatial coverage | Fixed | 4 soles | Increased spatial coverage |
|  | B | Secret Houses | Stochastic | 6 soles | None |
|  |  | Spatial coverage | Fixed | 2 soles | Increased spatial coverage |
|  | C | None (control) | n/a | n/a | None |
|  | D | Pay per detection | Stochastic | 40 soles | None |
| Cayma | A | Spatial coverage | Fixed | 2 soles | None |
|  |  | Risk information use | Fixed | * | Increased risk information use |
|  | B | Spatial coverage | Fixed | 2 soles | None |
|  |  | Secret Houses | Stochastic | 6 soles | None |
| JLByR<sup>2</sup> | A | Secret Houses | Stochastic | 10 soles | None* |
|  |  | Spatial coverage | Fixed | 1 sol | None |
|  | B | Pay per detection | Stochastic | 40 soles | None |
| Miraflores | A | Poker | Fixed | Vacation time (accrued in points) | Increased spatial coverage AND Increased risk information use |
|  | B | Pay per detection | Stochastic | ½ day off work | None |

### Socabaya trial

In this trial, there were six participants and four arms (A-D), which were distinguished by the incentive schemes tested: Secret houses, spatial coverage, and pay per detection. Over the course of four weeks, six participants visited a total of 2,824 houses and conducted 965 inspections in 24 search zones. Financial incentives significantly improved spatial coverage (Figure 3; Arm A: paired t-test, t = -5.4155, p < 0.01; Arm B: paired t-test, t = -7.6817, p < 0.001). The average size of the largest triangle in Arms A and B, (the two arms in which spatial coverage was incentivized) was 18.0 uninspected houses (range: 10.0 to 26.0) and 15.7 uninspected houses (range: 10.0 to 25.0), respectively. In Arms C and D, (in which spatial coverage was not incentivized), the largest group of uninspected houses averaged 52.7 (range: 35.0 to 71.0) and 50.0 (range: 35.0 – 72.0), respectively. We found no significant differences in risk information utilization in any of the arms (A, B and D) when compared with the control arm (Figure S1). The monetary value of the economic incentives awarded in this trial was an average of 10.1% of a participant’s salary (ranging from 4.0% to 13.0%).

**Figure 3.**
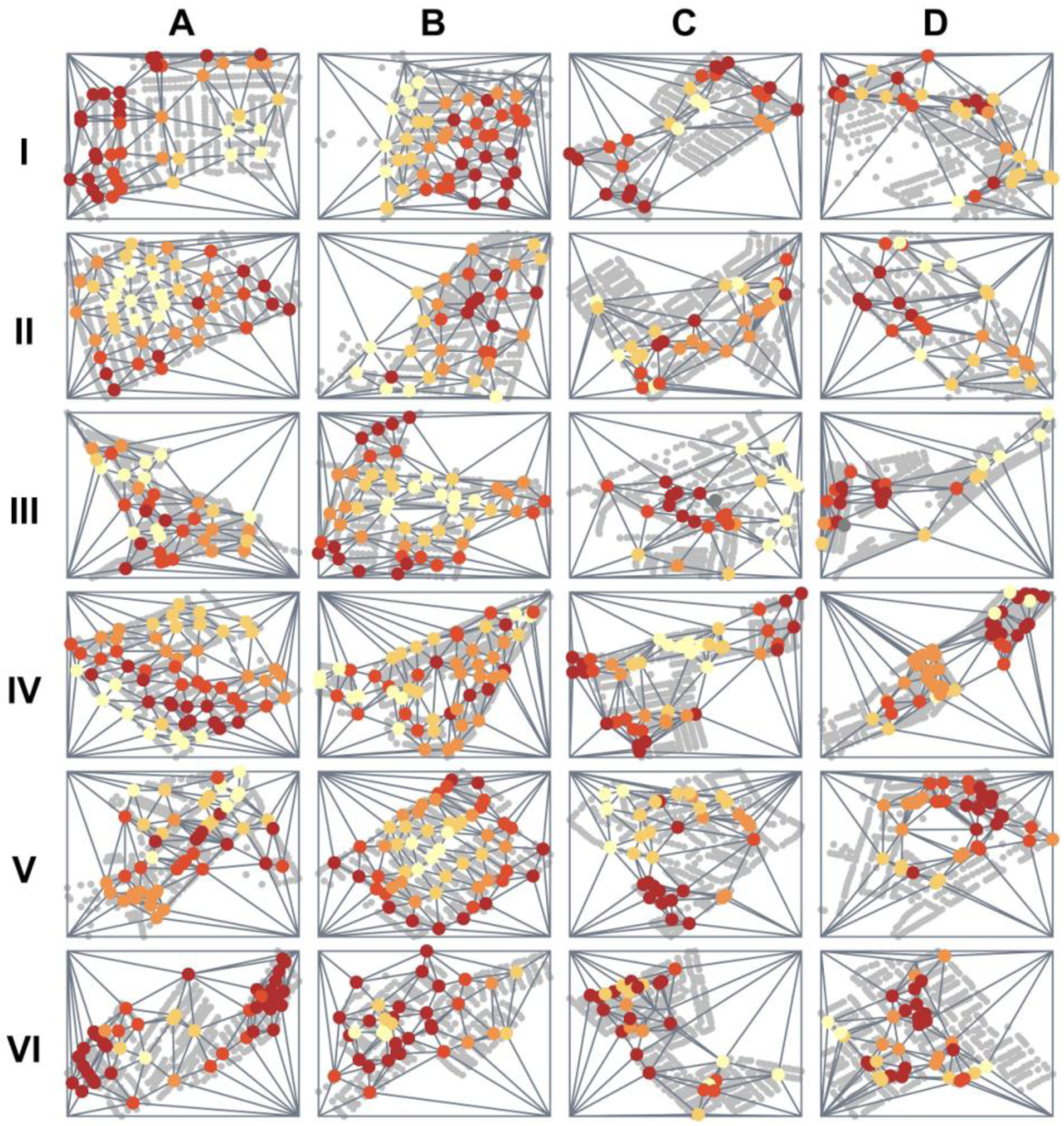
Spatial coverage maps for each participant in the Socabaya trial. Rows represent participants, columns represent trial arms. Incentives used in each arm were: (A) Fixed incentive for spatial coverage and a stochastic incentive for inspecting higher risk houses; (B) Fixed incentive for spatial coverage and an increased payout of the stochastic incentive for inspecting higher risk houses; (C) no incentives (control arm); and (D) pay per detection. Spatial coverage is represented by Delaunay triangulation in which triangles are formed connecting inspected houses (colored dots represent the inspected houses by the infestation risk shown as a gradient from yellow (lowest quintile) to dark red (highest quintile). Arms A and B both had significantly higher spatial coverage than Arms C and D (paired t-test, p < 0.01, p < 0.001, respectively).

### Cayma trial

We hypothesized that the relatively low use of risk information in the Socabaya trial might be due to the uncertainty involved in searching for secret houses, as compared to increasing spatial coverage. To test this hypothesis, we compared a fixed risk utilization incentive to the secret houses incentive. Here, there were six participants and two arms (A and B). For the fixed risk utilization, five reward amounts (0.1, 0.2, 0.3, 0.4 and 0.5 soles) corresponding to the five house infestation risk levels (lowest, low, medium, high, and highest) were awarded, as calculated by: 

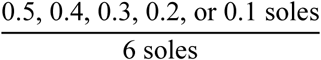

Six soles were selected for the denominator to align with the secret houses incentive amount in arm B.

The fixed payment significantly increased risk information utilization, (POLR model, OR 1.45, 95% CI [1.08 - 1.96], p < 0.02, Figure 4) compared to the stochastic payout scheme of the secret houses incentive. The average size of the largest triangle in arms A and B was 19.2 (range:10.0 – 44.0) and 17.8 (range: 9.0 – 29.0), respectively. There was no significant difference in spatial coverage between arms A and B. Payouts in arm A ranged from 1.0 % to 6.5% of the participants’ monthly salaries (mean 4.8%), and arm B payouts averaged 5.0% of a participant’s salary (ranging from 0.5% to 6.5%).

**Figure 4.**
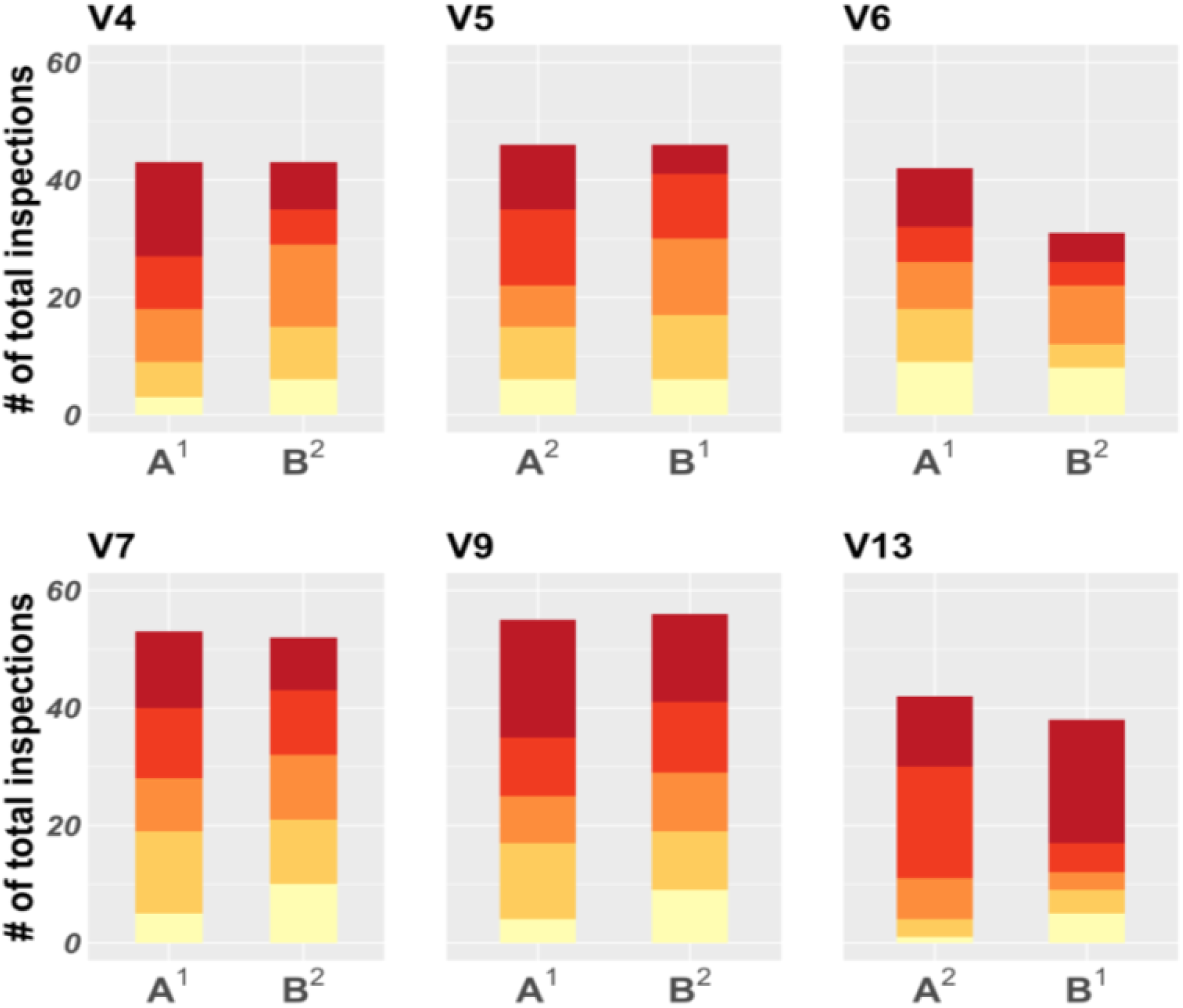
Infestation risk quintile distribution of households inspected by participants in the Cayma trial. Each set of two bars represents one participant, and each bar a study arm (A or B). A fixed incentive was used in arm A while the incentive in arm B was stochastic. Colors are ordered by risk quintile, going from the lowest (light yellow, bottom) to the highest (dark red, top). Arm A, (fixed incentive) had significantly higher risk information utilization than Arm B (stochastic incentive; POLR model, p < 0.02). Superscripts 1 and 2 in the x axis text indicate arm order.

### Jose Luis Bustamante y Rivero (JLByR) Trial

In this trial, we attempted to further incentivize utilization of the risk information in the maps by increasing the payout for inspecting higher-risk houses. There were six participants and two arms.

We had divergent results among participants in this trial. In arm A, one participant carried out 95% of their inspections in houses in the highest risk category, and the remaining 5% of their inspections were carried out in houses in the second-highest risk category, Figure 5). This participant greatly increased their use of risk information relative to arm B, but at the expense of spatial coverage. The other five participants did not significantly increase their inspections of higher risk houses in Arm A when compared to Arm B (POLR, odds ratio (OR) = 1.35, 95% CI [0.92-1.98], p < 0.2). The average size of the largest triangle in arms A and B was 42.5 (range: 10.0 – 120.0) and 60.0 (range: 20.0 – 77.0), respectively. We did not find any significant difference in spatial coverage between Arm A and Arm B. The average award in arm A was 0.67% of participants’ salaries (ranging from 0 to 1.71%).

**Figure 5.**
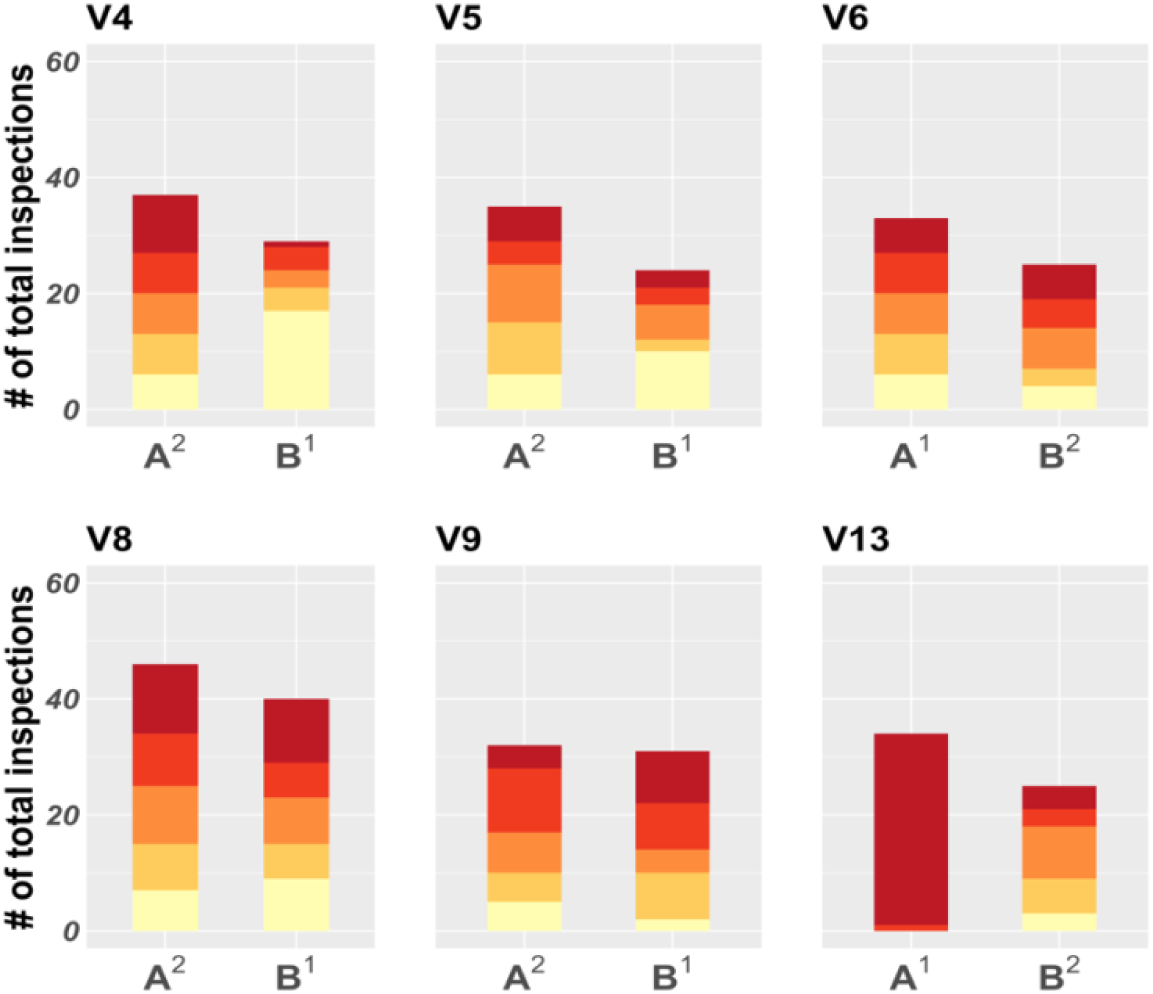
Distribution of household infestation risk quintile of households inspected by participants in the Jose Luis Bustamante y Rivero (JLByR) Trial. Each set of two bars represents one participant, and each bar, a study arm (A or B). Colors are ordered by risk quintile, going from the lowest (light yellow, bottom) to the highest (dark red, top). In arm A, the secret houses and spatial coverage incentives were used, while arm B employed only the pay-per-detection incentive. Superscripts 1 and 2 in the x axis text indicate arm order.

### Miraflores Trial

In arm A of this trial, we introduced a new incentive structure that was analogous to different hands of poker (described in detail in the Methods and in Figure 2). Rewards were given in the form of points that could be traded in for time off work, rather than monetary incentives. In arm B of this trial, we used the pay per detection incentive, with 500 points awarded for detecting an infested house. This trial had nine participants and two arms.

The poker incentive scheme resulted in achieving our dual objective of increasing risk information utilization (Figure 6) and spatial coverage of the search zone (Figure 7). Participants inspected a significantly greater number of high-risk houses (POLR, OR = 2.11, 95% CI [1.52-2.93], p < 0.001), and achieved better spatial coverage (paired t-test, t = 2.40, p < 0.05) when compared to the pay per detection arm. The largest group of uninspected houses averaged for poker arm and control arm contained 33.0 houses (range: 17.0 – 46.0) and 68.3 houses (range: 30.0 -109.0), respectively).

**Figure 6.**
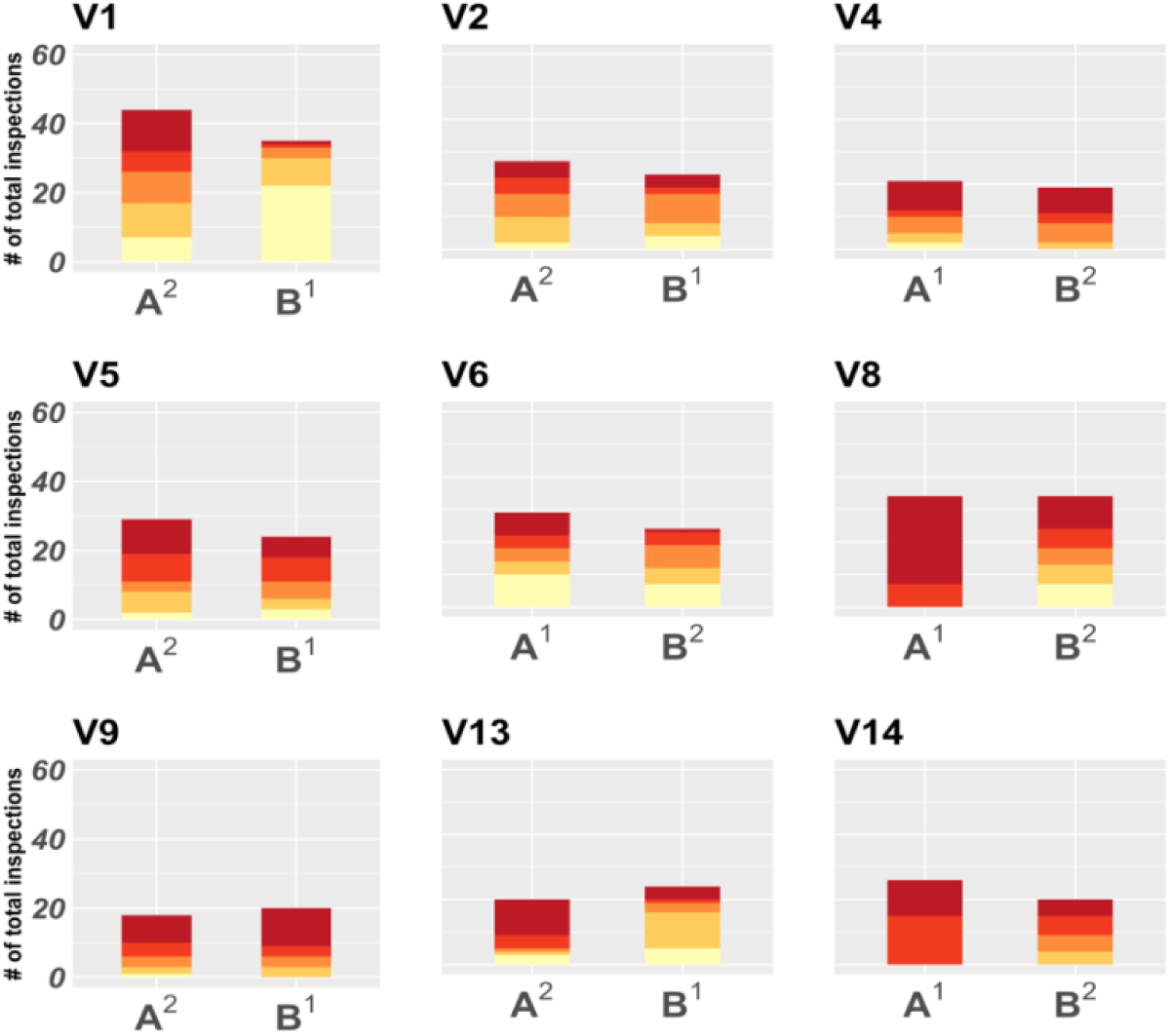
Distribution of household infestation risk quintile of households inspected by participants in the Miraflores Trial. Each set of two bars represents one participant, and each bar, a study arm (A and B). Colors are ordered by risk quintile, going from the lowest (light yellow, bottom) to the highest (dark red, top). Arm A was the poker incentive while arm B was pay per detection. The poker arm (A) had significantly different risk information utilization than arm B (POLR model, p < 0.001). Superscripts indicate arm order.

**Figure 7.**
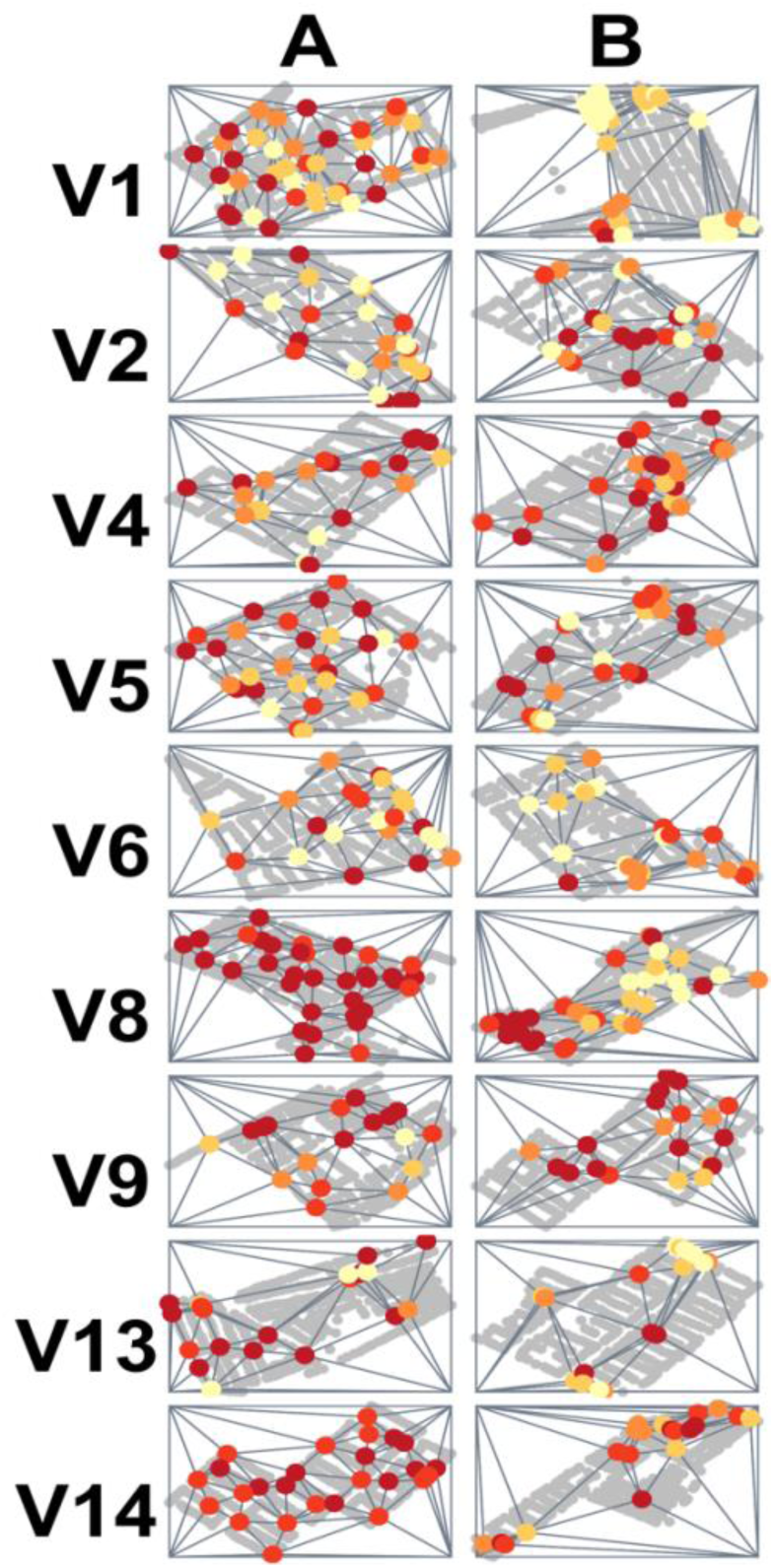
Spatial coverage maps for each participant in the Miraflores trial. Rows represent nine participants; columns represent trial arms. Incentives used in each arm were: A) poker incentive; and B) pay per detection. Spatial coverage is represented by Delaunay triangulation in which triangles are formed connecting inspected houses (colored dots represent the inspected houses by the infestation risk shown as a gradient from yellow (lowest quintile) to dark red (highest quintile). Arm A had significantly higher spatial coverage than Arm B (paired t-test, p < 0.05).

Two participants received half a day off (valued at 1.6 % of their monthly salary) by earning 500 points (V8 and V9) under the poker arm. No participant received a reward under the points per detection arm.

## DISCUSSION

The World Health Organization calls surveillance a ‘cornerstone’ of public health security and practice^22^, but if it is a cornerstone, it is one that must be constantly shaped to different architectures of control agencies, and new epidemiologic scenarios^23,24^. Urban disease vectors present a unique surveillance challenge, especially when infestations are infrequent and/or control personnel reduced. In these cases, evidence-based tools such as risk maps can enhance field surveillance, especially if used to their full advantage. In this series of rolling trials, we evaluated infestation risk map use by entomologic surveillance technicians under different incentive schemes. We used incentives to encourage the adoption of two search strategies: maximal spatial coverage of the surveillance zone and preferential inspections of houses with higher infestation risks. We found that technicians adopted one of the search strategies under several of the incentive structures, but our dual objective was only achieved under the poker scheme.

In pursuit of the incentive structure that would achieve our dual objective, we observed that stochastic incentives (secret houses and pay-per-detection) did not successfully influence participant behavior, even with payout amounts that were higher than those associated with fixed incentives. A preference for guaranteed payouts over lotteries or variable micropayments has been observed in some prior studies^25–28^, although many revolved around one-time executions of target behaviors^26–28^, and the collective findings are variable^29^. Regardless, a preference for the known versus the unknown (ambiguity aversion^30^) is a widely-observed phenomenon^31^, and it is fairly unsurprising that we observed this preference when incentivizing more complex behaviors.

Despite the success of fixed payments in incentivizing a single behavior, when we used weighted incentives to encourage both high spatial coverage and preferential inspections of higher risk houses, we failed. Participants apparently discovered that they could more easily maximize their earnings by focusing solely on the spatial triangulation. When incentives were more heavily weighted toward risk utilization, one inspector abandoned attempts to achieve high spatial coverage and focused solely on visiting higher risk houses^32^. This outcome may be due to the arduous nature of the work itself-searching for infrequent and sporadically distributed vectors across a large city is tedious at best. The simplest route to the fastest payout may well have been the more salient option if intrinsic motivation was low from the beginning, which has been observed in health-related behaviors (reviewed in^33–35^), although less so in health workers specifically^36^. Weighted incentives, at least in our case, seem to be limited in their ability to encourage complex behaviors. Only when we introduced the poker incentive scheme did participants both preferentially inspect higher risk houses and maximize their spatial coverage. In the poker scheme, the payouts for achieving each objective were complementary-there was no way to earn a higher reward without achieving both of them. For instance, to achieve the high scoring, “straight flush,” inspectors needed to both visit high risk houses and achieve substantial spatial coverage to receive a single fixed reward.

In epidemiologic surveillance, incentives or stipends have been used to increase data collection through new mobile tools, surveys, or disease testing, both at the individual/family level^37–40^ and at the community level through Community Health Workers (CHWs)^41–43^. Many recent studies tested uptake of new mobile disease surveillance tools in low and middle income countries^40,41,43,44^. A handful of vector control studies have successfully incentivized citizen scientists using cash rewards for data collection^45,46^, or by using paid, online crowd-sourcing to collect data^47^. Relatively few epidemiologic surveillance studies have attempted to use incentives to encourage more complex behaviors. In one notable study, salary incentives were used to increase uptake of a mobile, cloud-based tool for dengue surveillance by Public Health Inspectors in Sri Lanka. While spatial coverage was not encouraged with specific incentives, the authors observed that salary and bonus increases did result in a better spatial coverage^41^.

The poker scheme worked well for entomologic surveillance. As it is an easily communicated means to encourage multi-objective tasks, it could be generalized to other surveillance scenarios, and perhaps much more broadly. The success of the Miraflores trial also suggests that non-monetary incentives may be a feasible option when incentivizing complex behaviors, which may be especially useful for public health departments, whose resources are often scarce^48^. In addition to potentially being more cost-effective than cash payouts, some studies have found that non-monetary incentives have unique benefits such as increasing employee investment in their organization^49^, social reinforcement if other team members see the incentive received, and simply the enjoyment derived from looking forward to the incentive^50^.

Our trial was short for practical reasons; we do not know if we would obtain the same results over the years or decades-long course of a vector surveillance campaign. The poker incentive was used during the last study in a rolling trial, so it is possible that technicians became more accustomed to responding to incentives as they participated in each sequential study. Furthermore, we have previously found that there is a great deal of heterogeneity in surveillance among technicians, whether measured by sensitivity^51^ or productivity^9^. Given our relatively small sample size, individual differences could change in response to each incentive. Finally, we measured improvement in two domains: spatial coverage and risk level utilization. Although these metrics are used as surrogates for optimal infestation risk map use and effective surveillance, we do not know if improvement in both of these realms will lead to identifying more infested houses.

Eliminating disease vectors in cities involves familiar challenges: insecticide resistance, resistance to using insecticides in general, and resistance to changing the *status quo*^52,53^. In the post-spray phase of a largely successful vector control campaign, surveillance strategies must evolve and adopt new control methods that are suited to their post-spray scenario if they are to prevent vector re-emergence.

Implementation of data-based tools is a feasible option, although real improvements will only occur with the buy-in of those doing the day-to-day work in the field. Using an incentive such as the poker scheme, that is customized to achieve specific program objectives, is one potentially feasible approach to bridging the gap between tool design and optimal tool use.

## Data Availability

All relevant data are within the paper and its Supporting Information files. A public
repository with tools and code used in the app with the added triangulation feature is available in the following public repository: https://github.com/chirimacha/VectorPoint-Triangulation. Code for statistical analyses and relevant data for reproducibility is also located in the repository.

## Data Availability

All relevant data are within the paper and its Supporting Information files. A public repository with tools and code used in the app with the added triangulation feature is available in the following public repository: https://github.com/chirimacha/VectorPoint-Triangulation. Code for statistical analyses and relevant data for reproducibility are also located in the repository.

## ACKNOWLEDGEMENTS

The authors gratefully acknowledge the contributions of the Ministerio de Salud del Perú (MINSA), the Dirección General de Salud de las Personas (DGSP), the Estrategia Sanitaria Nacional de Prevención y Control de Enfermedades Metaxénicas y Otras Transmitidas por Vectores (ESNPCEMOTVS), the Dirección General de Salud Ambiental (DIGESA), the Gobierno Regional de Arequipa, the Gerencia Regional de Salud de Arequipa (GRSA), the Pan American Health Organization (PAHO/OPS), the Canadian International Development Agency (CIDA), Uptake Inc, Joseph Kable and Alexander Gutfraind.

## SUPPLEMENTARY MATERIAL

### S1. District descriptions

#### Socabaya

The district of Socabaya is located in Southwest Arequipa. The district has a geographic area of 18.64 km^2^ and a growing population of 80,000 people^21^. The insecticide spray portion of the vector control campaign took place in Socabaya in 2007, at which time *T. infestans* were detected in 9.8% of treated houses.

#### Cayma

The district of Cayma is located in the northern part of Arequipa, with a population of 103,458^54,55^ and a geographic area of 246 Km^2^. The insecticide spray phase of the *T. infestans* control campaign was carried out in Cayma in 2012, at which time 6.4% of targeted households in the district were infested (the district was sprayed in a focalized manner; not all the houses in the district were sprayed). Entomological surveillance has been more frequent in Cayma than in the Socabaya district.

#### Jose Luis Bustamante y Rivero (JLByR)

The district is located in southeast Arequipa, and borders Socabaya. JLByR has a geographic area of 11.06 Km^2^ with a population of 76,410 people^56^. The insecticide application portion of the vector control campaign in the JLByR district took place in 2006, at which time 12.5% of houses were found to be infested with *T. infestans*.

#### Miraflores

Located in Northeast Arequipa, this district has an area of 28.7 km^2^ and a population of 104,068 inhabitants^57^. The insecticide application phase of the vector control campaign was carried out in Miraflores in 2011, at which time 2.2% of houses were found to be infested with *T. infestans*.

**Table S1.** Spatial coverage data by participant per trial and arm (A-D). Columns contain the number of uninspected houses in the largest triangle formed by connecting the vertices of all inspected houses following a week of inspections. Dashes indicate that the participant did not participate in the trial. *Significant difference compared to control arm. Arm A: p < 0.01. Arm B: p < 0.001.

| Participant | Number of uninspected houses in the largest triangle |  |  |  |  |  |  |  |  |  |
| --- | --- | --- | --- | --- | --- | --- | --- | --- | --- | --- |
|  | Socabaya trial |  |  |  | Cayma trial |  | JLByR trial |  | Miraflores trial |  |
|  | A* | B* | C | D | A | B | A | B | A | B |
| V1 | - | - | - | - | - | - | - | - | 17 | 109 |
| V2 | - | - | - | - | - | - | - | - | 39 | 39 |
| V4 | - | - | - | - | 44 | 21 | 66 | 68 | 36 | 158 |
| V5 | 22 | 17 | 50 | 35 | 12 | 13 | 21 | 72 | 23 | 30 |
| V6 | 26 | 17 | 49 | 71 | 11 | 29 | 14 | 47 | 46 | 61 |
| V7 | 11 | 25 | 35 | 67 | 10 | 13 | - | - | - | - |
| V8 | 23 | 13 | 72 | 46 | - | - | 10 | 20 | 18 | 30 |
| V9 | 10 | 12 | 43 | 51 | 10 | 9 | 24 | 32 | 27 | 47 |
| V13 | 16 | 10 | 51 | 46 | 28 | 22 | 120 | 77 | 71 | 69 |
| V14 | - | - | - | - | - | - | - | - | 20 | 72 |
| MEAN | 18 | 15.7 | 50 | 52.7 | 19.2 | 17.8 | 42.5 | 60 | 33 | 68.3 |

**Table S2.** Monetary rewards earned by participants. Shown by trial, arm, and incentive type (risk information use and spatial coverage). Dash indicates no participation in the trial. Results from the final trial (Miraflores) are in Table 8, as a different payout type was awarded. Final row shows payout as a percentage of the inspector’s monthly salary (approximately $350 USD). ^1^One sol ≈ $0.28USD. *Significant difference in spatial coverage compared to the control arm (p < 0.001). ** Risk information use in the fixed arm was significantly different than in the stochastic arm (Cayma trial only; p < 0.02).

| Participant | Reward amount (in Peruvian soles<sup>1</sup>) |  |  |  |  |  |  |  |  |  |
| --- | --- | --- | --- | --- | --- | --- | --- | --- | --- | --- |
|  | Socabaya trial |  |  |  | Cayma trial |  |  |  | JLByR trial |  |
|  | A<br>(Stochastic) |  | B<br>(Stochastic) |  | A**<br>(Fixed) |  | B<br>(Stochastic) |  | A<br>(Stochastic) |  |
|  | *Risk info*<br>*use* | *\*Spatial*<br>*coverage* | *Risk info*<br>*use* | *\*Spatial*<br>*coverage* | *Risk info*<br>*use* | *Spatial*<br>*coverage* | *Risk info*<br>*use* | *Spatial*<br>*coverage* | *Risk info*<br>*use* | *Spatial*<br>*coverage* |
| V1 | - | - | - | - | - | - | - | - | - | - |
| V2 | - | - | - | - | - | - | - | - | - | - |
| V4 | - | - | - | - | 15.9 | 18 | 12 | 42 | 0 | 0 |
| V5 | 4 | 84 | 12 | 70 | 15.2 | 64 | 0 | 58 | 0 | 23 |
| V6 | 12 | 156 | 6 | 54 | 12.5 | 66 | 12 | 24 | 10 | 26 |
| V7 | 4 | 152 | 6 | 60 | 17.3 | 68 | 0 | 62 | - | - |
| V8 | 0 | 152 | 12 | 70 | - | - | - | - | 10 | 34 |
| V9 | 12 | 128 | 6 | 66 | 19.4 | 78 | 12 | 78 | 0 | 21 |
| V13 | 0 | 76 | 12 | 50 | 16.4 | 44 | 6 | 40 | 20 | 0 |
| V14 | - | - | - | - | - | - | - | - | - | - |
| Mean | 5.33 | 124.7 | 9 | 64.3 | 16.1 | 56.3 | 7 | 50.7 | 6.7 | 17.3 |
| % salary<br>(monthly) | 0.45% | 10.6% | 0.8% | 5.5% | 1.4% | 3.4% | 0.6% | 4.3% | 0.6% | 1.4% |

**Figure S1.**
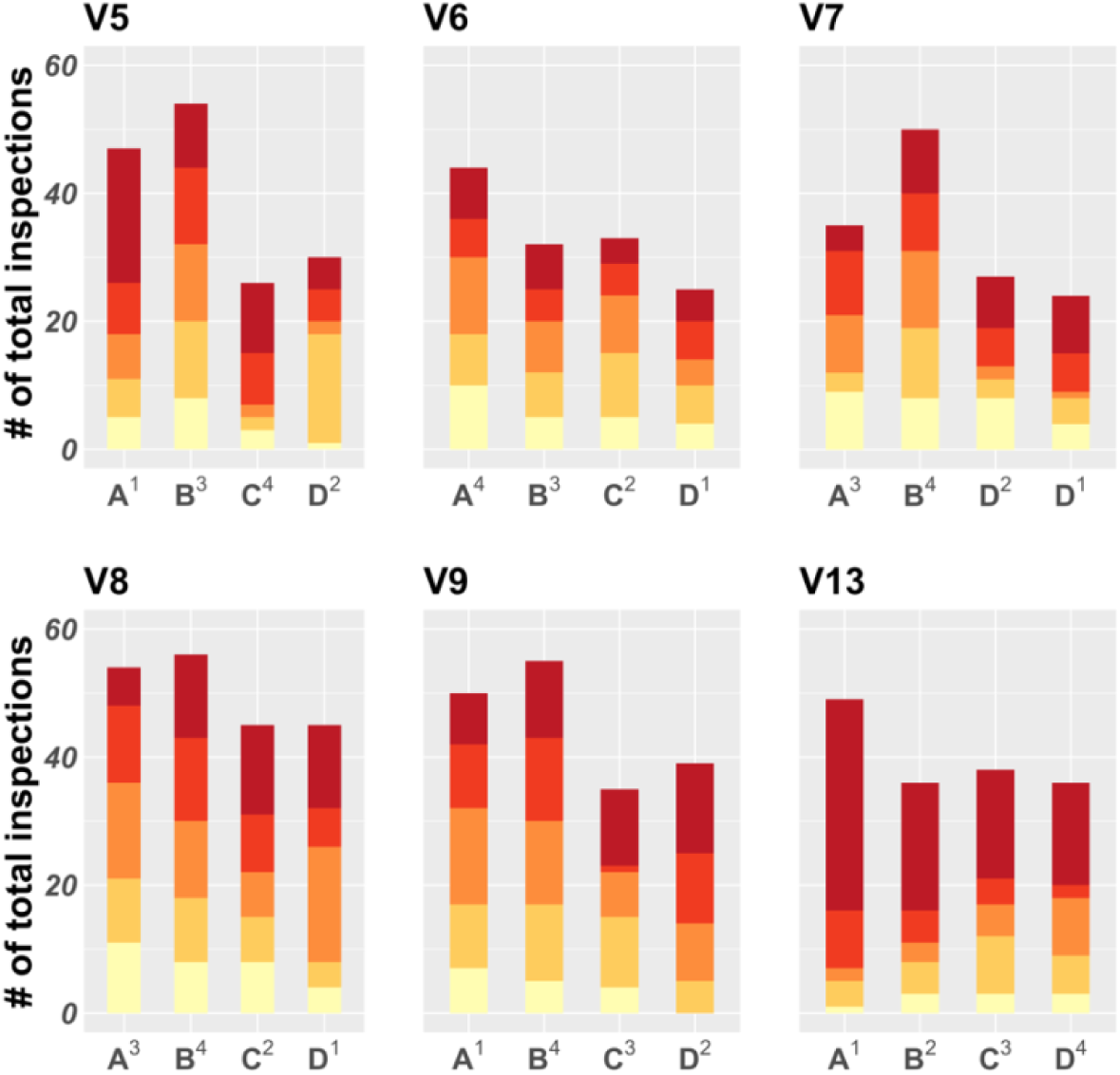
Distribution of infestation risk quintile for the households inspected by participants in the Socabaya Trial. Each set of four bars represents one participant, and each bar a study arm (A-D). Colors are ordered by risk quintile, going from the lowest (light yellow, bottom) to the highest (dark red, top). Superscripts above each arm name (A-D) in the x axis

